# Genetic modifiers of Huntington’s disease differentially influence motor and cognitive domains

**DOI:** 10.1101/2022.01.03.22268687

**Authors:** Jong-Min Lee, Yuan Huang, Michael Orth, Tammy Gillis, Jacqueline Siciliano, Eunpyo Hong, Jayalakshmi Srinidhi Mysore, Diane Lucente, Vanessa C. Wheeler, Ihn Sik Seong, Zachariah L. McLean, James A. Mills, Branduff McAllister, Sergey V. Lobanov, Thomas H. Massey, Marc Ciosi, G. Bernhard Landwehrmeyer, on behalf of the European Huntington’s Disease Network Registry investigators, Jane S. Paulsen, on behalf of the Huntington Study Group PREDICT-HD investigators, E. Ray Dorsey, Ira Shoulson, on behalf of the Huntington Study Group COHORT investigators, Cristina Sampaio, Darren G. Monckton, Seung Kwak, Peter Holmans, Lesley Jones, Marcy E. MacDonald, Jeffrey D. Long, James F. Gusella

**Author notes:** Correspondence (James F. Gusella). Principal investigators of the Genetic Modifiers of Huntington’s Disease (GeM-HD) Consortium. Clinical investigators contributing to the European Huntington’s Disease Network, Huntington Study Group and Enroll-HD studies are listed at links provided in the Acknowledgments.

## Abstract

Genome-wide association studies (GWAS) of Huntington’s disease (HD) have identified six DNA maintenance gene loci (among others) as modifiers and implicated a two step-mechanism of pathogenesis: somatic instability of the causative *HTT* CAG repeat with subsequent triggering of neuronal damage. The largest studies have been limited to HD individuals with a rater-estimated age at motor onset. To capitalize on the wealth of phenotypic data in several large HD natural history studies, we have performed algorithmic prediction using common motor and cognitive measures to predict age at other disease landmarks as additional phenotypes for GWAS. Combined with imputation using the Trans-Omics for Precision Medicine reference panel, predictions using integrated measures provided objective landmark phenotypes with greater power to detect most modifier loci. Importantly, substantial differences in the relative modifier signal across loci, highlighted by comparing common modifiers at *MSH3* and *FAN1*, revealed that individual modifier effects can act preferentially in the motor or cognitive domains. Individual components of the DNA maintenance modifier mechanisms may therefore act differentially on the neuronal circuits underlying the corresponding clinical measures. In addition, we identified new modifier effects at the *PMS1* and *PMS2* loci and implicated a potential new locus on chromosome 7. These findings indicate that broadened discovery and characterization of HD genetic modifiers based on additional quantitative or qualitative phenotypes offers not only the promise of in-human validated therapeutic targets, but also a route to dissecting the mechanisms and cell types involved in both the somatic instability and toxicity components of HD pathogenesis.

## Introduction

Huntington’s disease (HD [MIM: 143100]) is a dominantly inherited neurodegenerative disorder involving a constellation of motor, cognitive and behavioral manifestations caused by inheriting an expanded (> 35) CAG trinucleotide repeat in the coding sequence of *HTT* that lengthens the encoded polyglutamine segment in huntingtin.^1^ In sequential genome-wide association studies of HD^2-4^, we discovered that the timing of onset of characteristic HD motor signs (“age-at-onset”) is determined primarily by a property of the uninterrupted length of the inherited CAG repeat, not the encoded polyglutamine. The modification of age-at-onset by genetic variation at a number of DNA maintenance genes implicates length-dependent somatic instability of the CAG repeat segment. Moreover, in rare individuals with two expanded *HTT* repeats, age-at-onset is consistent with a dominant effect of the larger of the two CAG repeat alleles.^5^ Together, these findings support an HD process comprising at least two distinct components: 1) an initial phase in which somatic expansion of the CAG repeat to a critical threshold length (whose mechanism might act via the CAG repeat strand, the repeat CTG strand or both) determines the ultimate timing of disease onset; and, 2) a consequent phase in which neuronal toxicity, triggered in some manner by the somatically expanded repeat, produces progressive neuropathology and clinical manifestations, ending in early death of the individual.^4^ The driver of the neuronal toxicity remains uncertain. It could theoretically act at the DNA, RNA or protein levels, but those modifier associations that do not obviously relate to the process of somatic expansion do not point to a discrete toxicity mechanism.^3^ To date, the possibility for neuronal toxicity that has elicited the greatest experimental interest has been polyglutamine-mediated damage.^6^ Indeed, suppression of huntingtin expression in individuals with manifest HD is being actively pursued as an intervention strategy.^7; 8^ The discovery of genetic modifiers of HD age-at-onset established the proof-of-principle that the course of the disease can be altered prior to emergence of clinical manifestations, pointing to the process of somatic CAG expansion as the driver of the rate of onset and therefore as a therapeutic target to delay or prevent disease manifestations.

The potential for genetic modifiers of either of the two phases of HD pathogenesis to further elucidate the disease mechanism and to offer new routes for intervention, both before and after onset of manifestations, argues for further mining of the existing GWA data. The phenotype used in our previous HD GWAS was ‘residual age at motor onset’, based on the heritable variance in age-at-onset that remains after accounting for the contribution of inherited CAG repeat length.^2; 3; 5; 9; 10^ Age at motor onset was chosen for GWAS as the most frequently captured clinical measure available to maximize the sample size achievable by combining collections of HD research participants from historical genetic studies^1; 10; 11^ and from past or ongoing clinical trial and natural history studies.^12-16^ In the historical collections, age at motor onset was often the only disease landmark available. In the later clinical cohort studies, typically the Unified Huntington Disease Rating Scale (UHDRS)^17^ was used to capture an onset age along with a wide range of phenotypic measures across motor, cognitive, behavioral, and functional domains, collected cross-sectionally or longitudinally at different time points along the course of the disease. However, age specifically at motor onset was not always recorded when other signs of disease were the first to emerge.^18^

The integration of multiple common clinical measures can provide a prognostic index for motor diagnosis and measures of disease progression.^19; 20^ Indeed, an integrated phenotype (composite UHDRS (cUHDRS))^21^ derived from measures commonly collected via the UHDRS (*i.e*., TMS-Total Motor Score, SDMT-Symbol Digit Modalities Test, SW-Stroop Word, TFC-Total Functional Capacity) has recently been proposed as an effective measure for clinical trials, most of which are carried out in early manifest HD. Consequently, to extend the HD GWAS beyond age-at-onset, we have capitalized on the deep phenotype datasets from several HD natural history studies to perform algorithmic prediction of age at additional disease landmarks for participants in our existing GWAS dataset. Initially, we predicted the age at 50% probability of achieving two clinically defined landmarks: Diagnostic Confidence Level 4 (DCL4; indicating unequivocal motor signs with ≥99% confidence of HD) and a Total Functional Capacity score of 6 (TFC6; indicative of the beginning of Stage 3 on the Shoulson-Fahn functional capacity rating scale^22^, corresponding to when the capacity to perform activities of daily living is substantially reduced) (Figure 1 illustrates relationships for (CAG)42). We also re-imputed the GWAS genotypes using data from the Trans-Omics for Precision Medicine (TOPMed) Program^23^ as a larger, more diverse reference panel than the previous Haplotype Reference Consortium (HRC) data.^24^ The pattern of GWAS findings with these age-at-landmark phenotypes prompted us to also investigate phenotypic extremes of TMS and SDMT and algorithmically predicted TMS and SDMT landmarks. Together, these analyses revealed that the genetic differences at HD modifier loci can have differential impact on phenotypes preferentially related to motor or cognitive manifestations, indicating that not all HD modifier alleles impact equally the neuronal networks contributing to these phenotypes.

**Figure 1.**
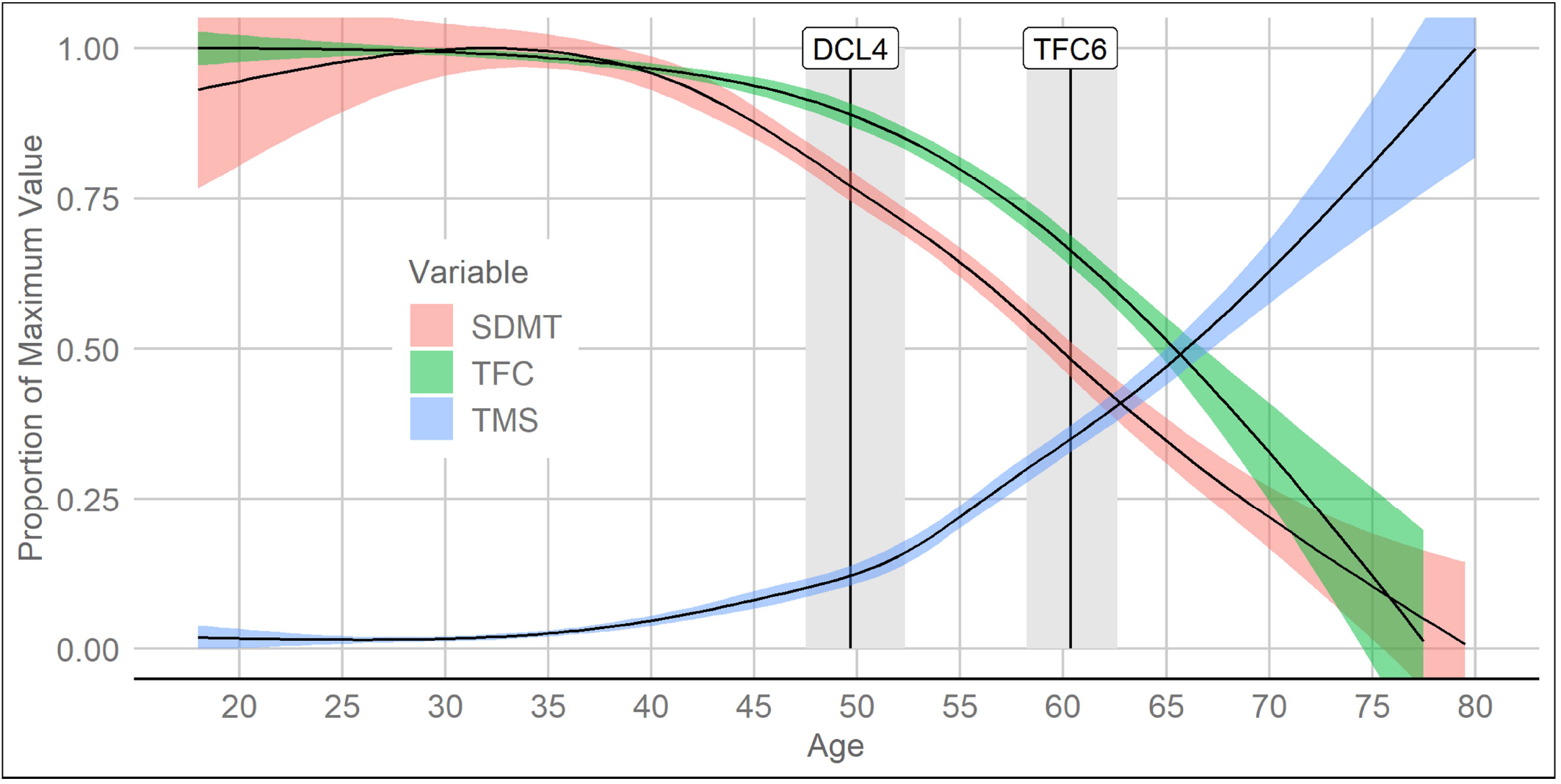
Trajectory of HD clinical measures using *HTT* CAG42 individuals as an example. Proportion of the maximum sample value of the UHDRS SDMT (red), TFC (green) and TMS (blue) by age for CAG = 42. The black lines running left to right are based on the predicted mean trajectories from linear mixed models with cubic splines of age and the colored ribbons are bootstrapped 95% confidence bands. Each curve value indicates the proportion of the maximum sample value observed for the variable in our dataset, with max(SDMT) = 110, max(TFC) = 13, and max(TMS) = 107. The vertical black lines denote the age on the horizontal axis at which there is estimated to be 50% conversion, respectively, to DCL4 and TFC6 (median survival), with 95% confidence intervals. Median survival (and its confidence interval) was estimated using a parametric survival model (Weibull regression) that accommodated left-truncation and right-censoring.

## Subjects and methods

### Subjects and clinical phenotypes

HD subjects in our modifier GWAS and genome-wide genotyping were described previously.^2; 3^ These genetic studies have been approved by the Institutional Review Board of Mass General Brigham (formerly Partners Healthcare). The phenotypic data were from the components of the UHDRS as recorded by trained study examiners in the PREDICT-HD study and the Cohort study of the Huntington Study Group, the Registry study of the European Huntington’s Disease Network and the ENROLL-HD platform. For the vast majority of participants, the age-at-onset of motor signs was that recorded by trained raters. For a small number of participants, a rater estimate was not available, so a family estimate was used. TMS, which can range from 0 (normal) to 124 (severest impairment), is the sum of items from the motor-assessment scale. This comprises the examiners’ evaluation of, among others, oculomotor function, dysarthria, chorea, dystonia, bradykinesia, gait, and postural stability. The language independent Symbol Digit Modalities Test (SDMT), adapted from the Wechsler digit symbol subtest, aims to measure working memory, complex scanning, and processing speed (but can also be influenced by motor dysfunction). Participants match symbols with numbers in a timed process whereby appropriate numbers are placed below the matching symbols as quickly as possible, generating raw scores based upon the number of items correctly completed in 90 seconds.

### Algorithmic prediction of age at HD landmarks

In our previous GWAS of HD onset, we relied on age at motor onset, which limits the sample size to those individuals who have passed a disease landmark whose assessment, by its nature, involves subjective judgment and, if judged retrospectively, potential recall bias. Consequently, we have explored the use of predictive modeling to generate GWAS phenotypes that integrate multiple individual standardized measures and reduce subjectivity to increase our power to detect associations. Computing a predicted phenotype for GWAS in HD is challenging. The number of individuals for which a phenotype is predicted should be maximized, while accounting for the vagaries of observational data. When the phenotype is a landmark event such as motor diagnosis, several difficulties present themselves. Participants can enter a study having experienced the event in the past, with the event timing being vague or unknown (left-censoring). Alternatively, individuals can enter the study and not experience the event in the time that they are observed (right-censoring). Visits tend to be annual and often irregular, so that the exact event timing can only be inferred to occur sometime between two visits (interval-censoring). An additional challenge is accounting for different ages at study entry when attempting to predict the age at which an event occurs. Age is the natural time metric for analysis with time 0 being birth. No participant has been observed at time 0 in an observational study, meaning that individuals have postponed or delayed entry (left-truncation), which requires proper identification of who is at risk of an event over what age interval. Finally, measurement of the event itself is tainted by error (*e.g*., natural waxing and waning of motor signs), and the amount of observed information (*e.g*., due to number of visits) among participants can vary widely. The above characteristics are very difficult to handle in a single statistical model. Therefore, for the primary predicted phenotypes of age-at-DCL4 and age-at-TFC6, we developed an algorithm for predicting age at each landmark event using both survival analysis and longitudinal analysis, described in Figure S1. We emphasize that the algorithm is intended to provide relatively accurate predictions for individuals in a manner that facilitates research and is not intended to make group-level inferences. From data on > 19,000 participants available through the Cohort, Predict-HD, Registry and Enroll-HD natural history studies, we predicted the age at each event and then utilized the subset of participants with available genome-wide genotyping data to perform association analyses.

In addition to age-at-DCL4 and age-at-TFC6, we defined additional landmarks based upon TMS or SDMT scores. We first determined the mean age-at-DCL4 and mean age-at-TFC6 for individuals at each CAG repeat length from 40 to 50 and compared these to the distribution of TMS and SDMT scores at each CAG length. For both TMS (scale: 0-107 in our sample) and SDMT (scale 0-110 in our sample), a score of 30 typically fell at an age between the median age at DCL4 and TFC6. Consequently, we selected this value for prediction of TMS and SDMT landmarks that occur within roughly the same age range of early manifest HD. Once the threshold value of 30 was decided, the estimated ages at which an individual progressed up to TMS = 30 (‘age-at-TMS30’) and down to SDMT = 30 (‘age-at-SDMT30’) was computed based on the third and fourth panels of Figure S1, except that linear mixed models for SDMT and TMS were used, respectively (rather than the model for cUHDRS from panel two).

### Selection of subjects for analysis of TMS and SDMT phenotypic extremes

TMS and SDMT phenotypes were classified as extreme as previously described.^25; 26^ In brief, TMS and SDMT collected as part of the UHDRS were classified as extreme if they were outside of predefined cut-offs on a distribution of that phenotype. Statistically, extremes of the phenotype distribution were derived from quantile regression (a linear quantile mixed model approach).^27^ For the longitudinal TMS and SDMT data, the quantitative phenotypes were classified as extreme at each visit if measures occurred outside of the cut-offs. The TMS data distribution was modelled as an exponential function of age stratified by CAG length. SDMT was examined on a linear function of age in ∼10-year age bins (18–29; 30–39; 40–49; 50–59; 60–69; 70–79; ≥ 80) controlling for CAG length and education. A single extreme measure may not mean the phenotype remains consistently extreme over time. Hence, participants were classified as being phenotypic extremes if they had an extreme phenotype, motor or cognitive, in the same direction at two or more consecutive visits. We defined the extremes as those below a 5% cut-off or above a 95% cut-off for TMS and below a 10% cut-off and above a 90% cut-off for SDMT in order to obtain roughly comparable numbers of genotyped subjects, since only a fraction of the study participants have genome-wide genotyping data.

### Imputation and association analyses

Five batches of GWA data sets were produced and subsequently imputed using the HRC reference panel as described previously.^3^ Genotype imputation using the TOPMed reference panel was performed similarly. Briefly, the individual genotype data set (*i.e*., GWA1, 2, 3, 4, and 5) comprising subjects with genotype call rate greater than 90% and single nucleotide variants (SNVs) with call rate > 95% and MAF > 1% was subjected to quality control by the “HRC or 1000G Imputation preparation and checking” program. Additional QC was performed using the TOPMed Imputation Server, revealing genomic regions of 10 MB with at least one sample with call rate < 50%. Those low call rate samples at certain genomic regions were further excluded from genotype imputation. The final QC-passed typed data set was used for imputation by the TOPMed Imputation Server. Genomic coordinates of TOPMed imputation data were converted to GRCh37/hg19 to make this data set directly comparable to the HRC imputation data set. Finally, we removed SNVs with imputation r-square < 0.5 in any of the GWA data sets, HWE p-value < 1E-6 (excluding chr4:1-5000000 region containing *HTT*), or MAF < 0.1%. For a given phenotype, genome-wide association analysis was performed for individuals of European ancestry using a mixed effects model with a relationship matrix and a set of covariates such as sex, batch, and the first four principal component values from genetic ancestry analysis as covariates.^3^ For selected candidate regions with significant association signals, conditional analysis was performed using a fixed effect linear model that included one or more additional covariates representing the minor allele counts of conditioned SNVs. SNVs are expressed as chromosome_coordinate_reference-allele_alternative-allele based on the GRCh37/hg19 genome assembly. CAG repeat lengths for each subject were generated by a standard ABI fragment analysis assay.^3; 28^ For individuals uncovered in the TopMed imputation with *HTT* CAA-loss and CAACAG-duplication haplotypes, whose CAG lengths are systematically mis-estimated by the fragment analysis assay, the pure CAG repeat size was determined by MiSeq (Illumina) DNA sequencing analysis using the previously reported assay method.^3^

### Gene-wide and Pathway Analysis

Gene-wide association analyses were carried out in MAGMA^29^ on summary statistics from each GWAS using genotypes from 7,086 individuals from GWA3, 4 and 5 as a reference panel to estimate LD. As in our previous HRC-based analysis^3^, we used a window of 35kb upstream and 10kb downstream of gene positions (GRCh37/hg19) along with the ‘‘multi’’ analysis option, combining the mean SNV p-value with the top SNV p-value (corrected for number of SNVs and the LD between them). Pathway enrichment analyses were also performed in MAGMA, correcting for LD between genes, initially using ‘‘self-contained analysis’’ measuring overall association among genes in a pathway, and then using a more conservative ‘‘competitive’’ analysis, to compare association in genes within a pathway to those outside the pathway.

### Pathway definition

The assignment of Gene Ontology (GO) terms to human genes was obtained from the “gene2go” file, downloaded from NCBI on March 11^th^, 2020. “Parent” GO terms were assigned to genes using the ontology file downloaded from the Gene Ontology website on the same date. GO terms were assigned to genes based on experimental or curated evidence, excluding electronic or computational annotations. Reactome pathways were downloaded on April 26^th^, 2020. Biocarta, KEGG and Pathway Interaction Database (PID) pathways were downloaded from v7.1 (March 2020) of the Molecular Signatures Database.^30^ Analysis was restricted to GO terms containing between 10 and 2000 genes. No size restrictions were placed on the other gene sets, since these were fewer in number. This resulted in a total of 10,041 gene sets for analysis.

### Statistics and software

TOPMed genotype imputation was based on the TOPMed Imputation Server. Mixed effects model GWAS was performed using the gemma program version 0.94 beta.^31^ Genomic inflation factors were calculated using the R package ‘GenABEL’ (version, 1.8-0).^32^ Estimated age-at-landmarks were developed using the R package flexsurv^33^ for survival analysis and lme4 for longitudinal linear mixed models.^34^ Quantiles for extreme values of the TMS and SDMT were determined using the R package lqmm.^27^ Gene-wide and pathway analyses were performed with MAGMA v1.08. Gene Ontology annotations were downloaded from the NCBI and Gene Ontology websites. Reactome pathways were downloaded from the Reactome site. Biocarta, KEGG and Pathway Interaction Database (PID) pathways were downloaded from v7.1 of the Molecular Signatures Database.

## Results

### TOPMed imputation-based age-at-onset GWAS

In our most recently reported HD age-at-onset GWAS, we analyzed 9,058 individuals of European ancestry whose genome-wide SNV data was imputed from array-typed genotypes using HRC data.^3^ This GWAS identified nine genome-wide significant loci tagged by SNVs of minor allele frequency (MAF) > 1%. Six of these loci harbor DNA maintenance genes (*FAN1, MLH1, MSH3, PMS1, PMS2, LIG1*) and, for three of these (*FAN1, MSH3, LIG1)*, we further defined multiple haplotypes representing distinct modifier effects.^3; 35^ The other three loci harbor candidate genes (*RRM2B, TCERG1, CCDC82*) that might affect somatic instability indirectly or, alternatively, might influence subsequent toxicity/response mechanisms. We also reported SNVs tagging two infrequent (< 1%) *HTT* haplotypes that differ from canonical CAG expanded alleles. In canonical alleles, the CAG repeat is followed by a single CAACAG codon doublet but in the variant alleles the CAA interruption is either lost or the CAACAG doublet is duplicated.^3; 36-38^ These variants cause the length of the uninterrupted CAG repeat to be estimated incorrectly in a standard PCR-based fragment-sizing assay using canonical sequence standards. The initially mis-estimated CAG length resulted in incorrect assignment of expected age-at-onset, which then produced false association signals. After correction of the mis-estimated CAG sizes by direct sequencing in those subjects identified by SNVs tagging the false signal, the CAACAG duplication haplotype was no longer significant while the CAA loss haplotype continued to yield signal at the border of genome-wide significance (p-value 5E-08). Finally, we also reported several additional loci detected only by single SNVs at < 1% MAF but suggested that these might be spurious due to outlier effects and required further confirmation.

For the present study, we re-imputed genotypes genome-wide using TOPMed data^23^ as a reference set and again performed GWAS of residual age-at-onset as a continuous phenotype for those 9,009 individuals whose data passed quality control. In preliminary analyses, a series of variants flanking the *HTT* CAG repeat location identified additional individuals carrying the rare CAA-loss and CAACAG-duplication haplotypes who had been missed by HRC imputation. We sequenced their CAG repeat region to correct the CAG repeat lengths and corresponding age-at-onset residuals prior to repeating a GWAS that yielded results largely comparable to HRC imputation for the 9,009 individuals (Figure S2). However, in the relationship between CAG repeat length and age-at-onset, the range of residual age-at-onset values is greater at lower than at higher repeat length (Figure S3). Consequently, to allow direct comparison with the Z score-based algorithmically-predicted landmarks (below), we calculated Z-scores for age-at-onset at each individual CAG repeat length for use as the continuous phenotype in GWAS.

The TOPMed-based Z-score age-at-onset GWAS (Figure 2) detected all of the loci previously implicated at genome-wide significance by SNVs of > 1% MAF except the chromosome (chr) 11 *CCDC82* locus, which in our previous GWAS was detected at genome-wide significance only by a dichotomous analysis of allele frequencies in the 30% extremes of residual age-at-onset.^3^ At *MSH3, FAN1*, and *LIG1*, where we have reported multiple modifier effects, SNVs tagging each of these effects again showed at least suggestive evidence of association (p-value < 1E-05) with age-at-onset (Table S1). With one exception, those peak SNVs in the current analysis that were not identical to the previously reported tag SNVs showed similar allele frequency, direction of effect and strong allele correlation (r-square > 0.97) with the prior tag.^3^ The peak SNV associated with onset-delay at the *PMS2* locus showed a slightly lower correlation with the previously assigned 7AM1 tag SNV (r-square = 0.86). In addition, second modifier effects (here termed 2AM2 and 7AM2) were detected in the vicinity of *PMS1* (p-value 3.67E-06 at 2_190639862_C_T) and *PMS2* (p-value 2.44E-07 at 7_6056484_G_C) based upon suggestive significance of SNVs associated with onset-delaying and onset-hastening modification, respectively, opposite in direction to the 2AM1 and 7AM1 modifier effects. Notably, after the correction for uninterrupted CAG repeat length, the rare CAA-loss and CAACAG-duplication *HTT*-region haplotypes continued to support genome-wide and suggestive significant association, respectively (Figure S4, Table S1). This suggests either that these changes to the CAG repeat region sequence alter its inherent propensity for somatic instability, changing the rate of CAG expansion leading to onset, that they influence the later toxicity mechanism, or that there is a separate linked modifier variant in 4p16.3 on one or both of these extended haplotypes.^3; 38^ In addition to previously recognized loci, a novel genome-wide significant onset-delaying signal emerged on chr 7 (7BM1: p-value 7.29E-09 at 7_56241504_C_T; Figure 2; Table S1) near the testis-expressed *NUPR2* (nuclear protein 2, transcriptional regulator), and upstream of both *CHCHD2* (coiled-coil-helix-coiled-coil-helix domain containing 2), which encodes a negative regulator of apoptosis involved in response to mitochondrial stress, and *PHKG1* (phosphorylase kinase catalytic subunit gamma 1), which regulates glycogenolysis. Finally, genome-wide analysis of SNVs with very low MAF (> 0.1% but < 1%) failed to reproduce any of the ‘hits’ represented by single rare SNVs reported previously from HRC imputation,^3^ reinforcing the need for caution in interpreting such loci.

**Figure 2.**
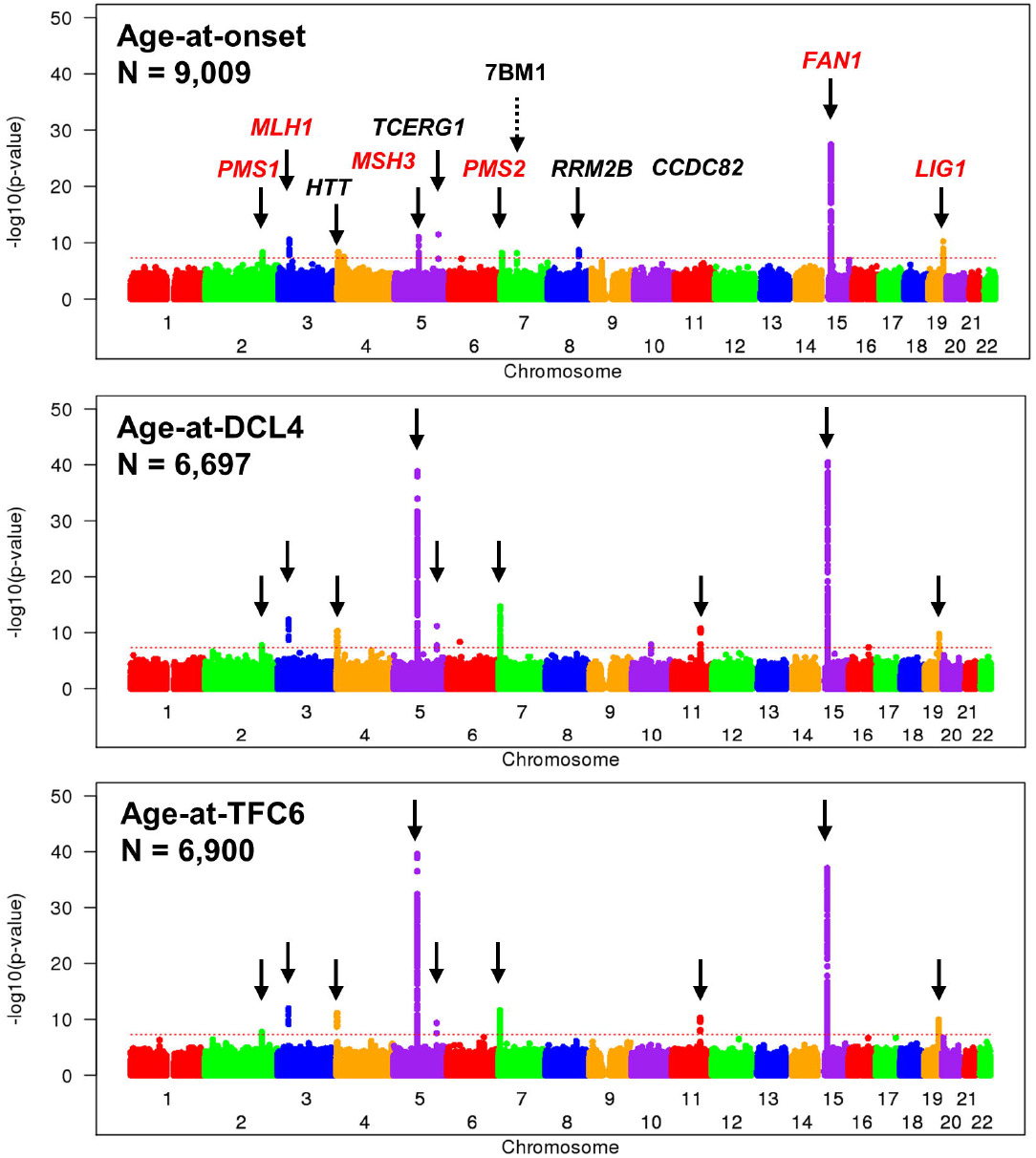
GWAS of age-at-onset, age-at-DCL4 and age-at-TFC6. Stacked plots comparing GWAS are shown for the Z score of CAG-specific age-at-onset, the Z score from age-at-DCL4 prediction and the Z score from age-at-TFC6 prediction. Solid arrows indicate loci that yield genome-wide significant signal with SNV of > 1% MAF (with the exception of *HTT*), with candidate modifier genes all labeled in the top panel. One novel locus that awaits further confirmation is noted by a dashed arrow and its HD modifier effect designation (7BM1) based upon our standard nomenclature.^3^ Red labels indicate DNA maintenance genes.

### Algorithmic prediction and GWAS of additional HD landmarks

To perform GWAS for additional landmarks during HD pathogenesis, we explored the use of predictive modeling, as illustrated in Figure S1, to produce estimates of the ages at which each individual had a 50% probability of reaching DCL4 and TFC6, respectively. Briefly, we generated a survival curve for the landmark phenotype (either DCL4 or TFC6) for each combination of CAG repeat length, sex, and education level (low = high school or less, or high = more than high school) to determine the median age at survival (*i.e*., the age at which 50% of the individuals have reached the landmark phenotype). We also modeled the mean trajectory for the cUHDRS to determine the cUHDRS value at the aforementioned median survival age. Individual cUHDRS trajectories extrapolated over the entire age range then predicted the individual’s age that corresponded which this predetermined cUHDRS value. The individual age-at-landmark prediction was then transformed to a *Z*-score by correcting for the median age and scaling by the standard deviation (SD) of all the ages for the individuals in the same CAG, sex and education cohort.

The landmark predictions were made using longitudinal clinical data from 19,173 participants in the Predict-HD, COHORT, Registry and Enroll-HD studies. Genome-wide genotyping data are currently available for ∼7,400 of these participants, ∼5,200 of which could be analyzed in the age-at-onset GWAS. The predicted age-at-DCL4 and age-at-TFC6 phenotype Z-scores were highly correlated with each other and both were less correlated with the age-at-onset Z-score (Table 1). We tested the capacity of these phenotypes to capture modifier loci by age-at-landmark GWAS directly comparable to the age-at-onset Z-score analysis. As shown in Figure 2 and in Table S1, despite the smaller sample size for both age-at-DCL4 (N = 6,697) and age-at-TFC6 (N = 6,900) landmarks compared to age-at-onset (N = 9,009), the predicted phenotypes revealed genome-wide significant signals at all but one of the previously reported loci.

**Table 1.** Correlation between Landmark Phenotypes*.

|  | Age-at-Onset Z-score | Age-at-DCL4 Z-score | Age-at-TFC6 Z-Score | Age-at-TMS30 Z-score | Age-at-SDMT30 Z-score |
| --- | --- | --- | --- | --- | --- |
| Age-at-Onset Z-score |  | 0.47 | 0.47 | 0.44 | 0.24 |
| Age-at-DCL4 Z-score |  |  | 0.97 | 0.81 | 0.74 |
| Age-at-TFC6 Z-Score |  |  |  | 0.83 | 0.75 |
| Age-at-TMS30 Z-score |  |  |  |  | 0.56 |

\* Pearson correlation coefficient based upon the 4,879 individuals with all five phenotypes

Two intriguing aspects of the age-at-landmark GWAS were the striking differences compared to the age-at-onset GWAS in the level of significance at some loci and the changes in their relative significance compared to other loci in the same analysis. The modifier signals for some loci, such as *MSH3, PMS2, CCDC82* and *FAN1* were increased substantially, while others, such as *PMS1, TCERG1, RRM2B* and *LIG1* were not. For most loci, the source of the signal could be equated confidently to the same modifier effect captured by age-at-onset because the peak signal was attributed to the same SNV or to one very highly correlated with it (r-square > 0.97), although exceptions at *MSH3, PMS2* and *LIG1* hinted at a potentially greater complexity underlying these modifier effects (Table S1).

The change in relative signal was most dramatic at *MSH3*. In the age-at-onset GWAS, the common 15AM2 delaying modifier at *FAN1* was far more significant (peak p-value 7.23E-28 at 15_31241346_G_A) than the common 5AM1 hastening modifier at *MSH3* (peak p-value 1.05E-11 at 5_79913275_G_A). By contrast, while both 5AM1 and 15AM2 showed greater significance in the age-at-DCL4 and age-at-TFC6 analyses than the maximum achieved in the larger age-at-onset sample, the signal for 5AM1 was disproportionately higher (peak p-values 2.69E-40 and 1.42E-39, respectively, at 5_79950781_A_G), becoming comparable to 15AM2 (peak p-values 9.03E-38 and 3.45E-41, respectively, at 15_31241346_G_A). This change in absolute and relative signals was not due to differences in the sample size or in the individuals tested, since analysis of only those 5,009 individuals where all three phenotypes were available generated similar results (5AM1: peak p-values 9.18E-07 for age-at-onset, 6.26E-32 for age-at-DCL4 and 1.06E-33 for age-at-TFC6, at 5_79950781_A_G; 15AM2: peak p-values 8.55E-14 for age-at-onset, 5.33E-33 for age-at-DCL4, 8.48E-34 for age-at-TFC6, at 15_31241346_G_A).

Relative p-value improvement in GWAS of the cUHDRS-predicted landmarks was also particularly notable at *PMS2* (Figure 2), again indicating that the age-at-onset and predicted landmark phenotypes had a differential capacity to reveal effects of particular modifiers. In this instance, the top SNV associated with landmark delay (7_6041836_T_A) had a much higher MAF than the 7AM1 age-at-onset modifier tag (MAF ∼41% vs. ∼15%), pointing to an additional modifier effect and thereby prompting the naming of a third modifier effect (7AM3) at the locus (Table S1).

The relative p-value improvement with the algorithmically predicted landmarks was also evident beyond the DNA maintenance genes. Both age-at-DCL4 and age-at-TFC6 robustly detected the *CCDC82* locus at far greater significance than age-at-onset. The *HTT* CAA-loss and CAACAG-duplication haplotypes also showed increased significance in the predicted-landmark GWAS. Surprisingly, the predicted landmarks yielded only suggestive significant signal at *RRM2B*, a locus that emerged as genome-wide significant in our first HD GWAS^2^ and was confirmed in our most recent larger GWAS^3^ (as well as in the age-at-onset analysis in Figure 2). Overall, the age-at-DCL4 and age-at-TFC6 analyses appeared to track a subset of established modifier loci and did not uncover evidence for a frequent modifier that acts after the DCL4 landmark by a new mechanism. However, along with the age-at-onset GWAS, the age-at-DCL4 and age-at-TFC6 GWAS did nominate several new loci represented by alleles at MAF < 1% (Table S2) that, like the new chr7 locus seen only in the age-at-onset analysis, will require future confirmation as *bona fide* modifiers since each could result from a small number of phenotypic outliers.

### Dichotomous analysis of TMS and SDMT extremes

The selectively greater capacity of the predicted landmarks to detect particular loci, such as *MSH3* and *PMS2*, relative to other known age-at-onset modifiers pointed to a difference in the nature of these phenotypes compared to the observed age-at-onset phenotype. Age-at-onset was typically determined by expert raters based upon motor signs characteristic of HD whereas the algorithmic prediction of age-at-DCL4 and age-at-TFC6 made use of motor, cognitive and functional measures in the cUHDRS in refining the age-at-landmark predictions. Furthermore, the age-at-onset analysis was limited to individuals who had the event of onset, whereas the prediction-based analysis had no such constraint. Consequently, we reasoned that, as opposed to landmarks that reflect a combination of motor and cognitive impairment, a direct comparison of two measures with no constraining event, the TMS and SDMT, aimed primarily (though not exclusively) at motor and cognitive domains, respectively, might reveal a differential impact of the modifiers on these domains.

In an initial comparison, we examined the longitudinal records of the Registry study of the European Huntington’s Disease Network and of the Enroll-HD study and identified those individuals who, for a given CAG repeat length, consistently fell within the extremes of the TMS or SDMT score distributions. We then performed a dichotomous GWAS comparing SNV allele frequencies between those extreme high and low scoring individuals for whom genome-wide genotyping data were available (for TMS (N = 920) and for SDMT (N = 1,200)) (Figure 3). The *FAN1* locus showed genome-wide significance for both phenotypes with peak-scoring SNVs being associated with lower TMS and higher SDMT score (top SNV p-value TMS: 4.54E-17 at 15_31241346_G_A; SDMT: 1.23E-12 at 15_31279883_CTTTT_C) indicative of less disruption of both motor and cognitive domains. By contrast, the *MSH3* locus failed to show even suggestive significance for the TMS phenotype but yielded two distinct peak signals in the SDMT extreme analysis for SNV’s whose minor alleles were associated with better and worse performance, respectively (top SNV p-values 6.64E-16 at 5_79915020_G_A and 8.54E-15 at 5_79947431_A_G). No other locus showed genome-wide significance in either of these small, highly selected TMS-and SDMT-extreme samples.

**Figure 3.**
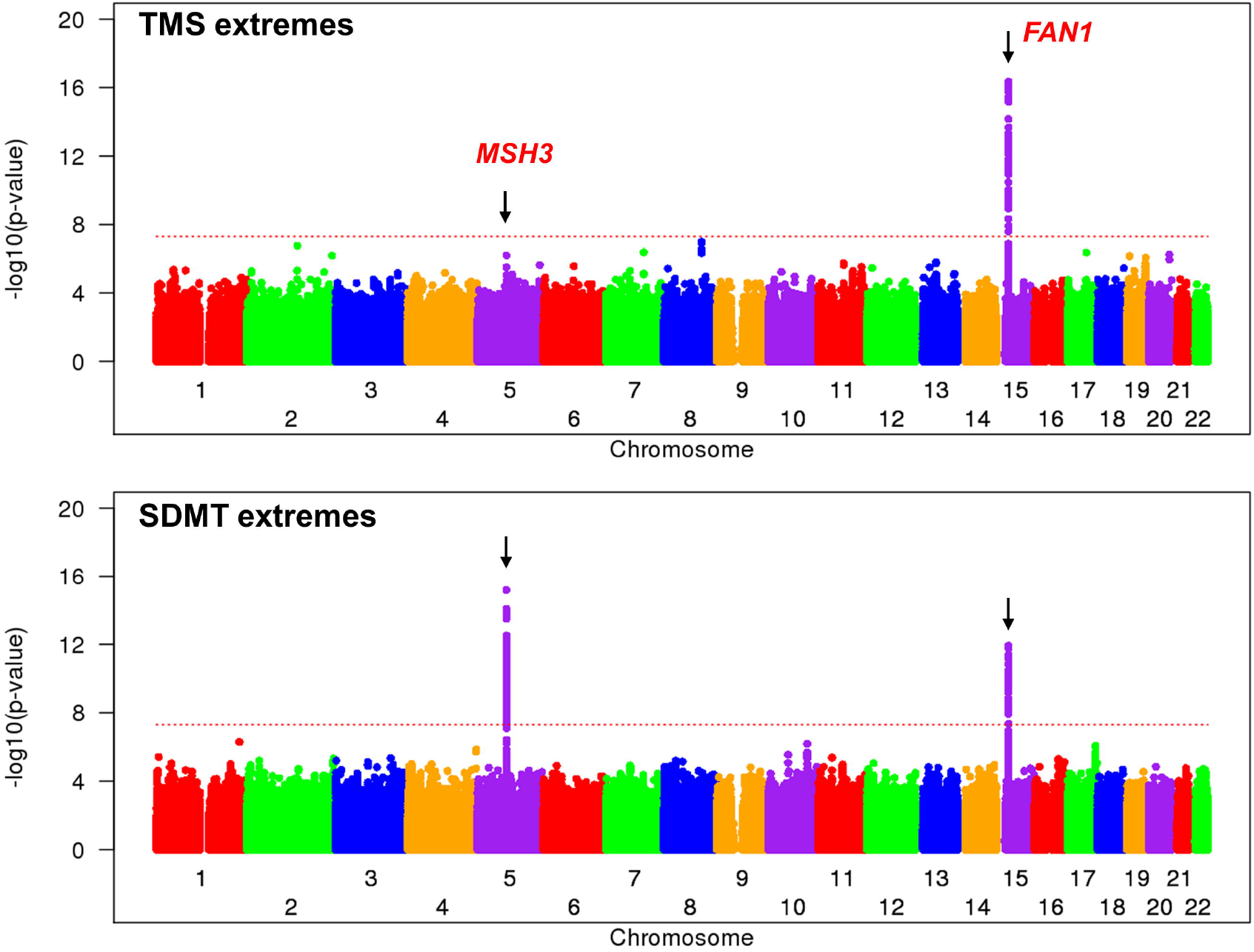
Dichotomous GWAS of Total Motor Score and Symbol Digit Modalities Test extremes. Stacked plots of dichotomous GWAS comparing allele frequencies among HD subjects in the Registry study who represented consistent phenotypic extremes with respect to TMS (top) or SDMT (bottom). Arrows mark the two loci that yielded genome-wide significant signal in at least one of the analyses with modifier gene labels denoted in the top panel.

### Analysis of TMS and SDMT score landmarks

To address the differential action of the modifier loci on TMS and SDMT in a manner directly comparable to our age-at-landmark predictions of DCL4 and TFC6, we then used algorithmic prediction similar to Figure S1 panels 3 and 4 (using linear mixed models for SDMT and TMS rather than cUHDRS) to compute estimated ages in early manifest HD at which each individual progressed up to TMS = 30 (N = 6,897) and down to SDMT = 30 (N = 6,727). The resulting age-at-TMS30 and age-at-SDMT30 landmark Z-scores were more highly correlated with the age-at-DCL4 and age-at-TFC6 Z scores than with each other or with the Z-score for age-at-onset (Table 1).

The GWAS for these two new landmark phenotypes (Figure 4 and Table S1) reinforced the capacity of distinct disease phenotypes to detect individual modifier loci differentially. For example, the *MSH3* locus showed relatively greater significance than *FAN1* for age-at-SDMT30. The relative differences in modifier peaks for age-at-TMS30 and age-at-SDMT30 were not due to the slightly different samples tested, since p-values calculated based upon only the 6,716 participants who were scored for both age-at-TMS30 and age-at-SDMT30 did not substantially alter the results (*e.g*., joint set vs. full sample p-values for 5_79950781_A_G (5AM1): age-at-TMS30 [3.62E-19 vs 1.35E-23] and age-at-SDMT30 [4.96E-37 vs 4.70E-37]; for 15_31241346_G_A (15AM2): age-at-TMS30 [2.51E-41 vs 7.41E-45] and age-at-SDMT30 [7.19E-22 vs 1.34E-21]. As expected, the different information content of these landmark phenotypes was also evident in conditional analyses, where the age-at-TFC6 signal at *MSH3* was preferentially reduced by conditioning on age-at-SDMT30 whereas at *FAN1*, it was preferentially reduced by conditioning on age-at-TMS30 (Figure 5). Taken together with the SDMT extreme analysis, these results confirmed that the integration of SDMT into the cUHDRS, which was used in the prediction of age-at-DCL4 and age-at-TFC6, at least partially explains the relatively increased *MSH3* signal in those GWAS compared to that for age-at-onset.

**Figure 4.**
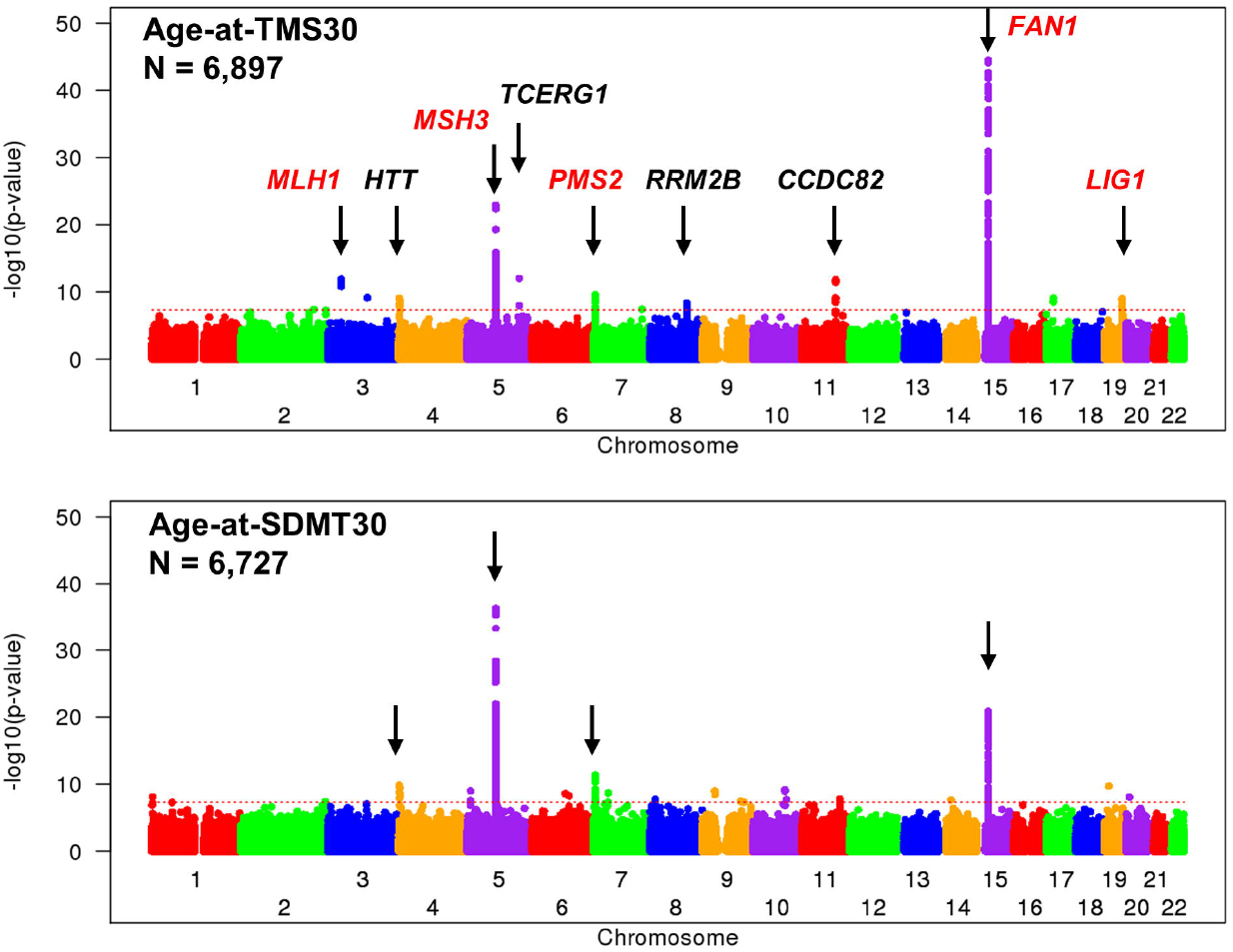
GWAS of Total Motor Score and Symbol Digit Modalities Test landmarks. Stacked plots comparing GWAS analysis of age-at-TMS30 and age-at-SDMT30. Arrows indicate loci that yielded genome-wide significant signal for variants of >1% MAF (with the exception of *HTT*), with the candidate genes labeled in the top panel. Red labels indicate DNA maintenance genes.

**Figure 5.**
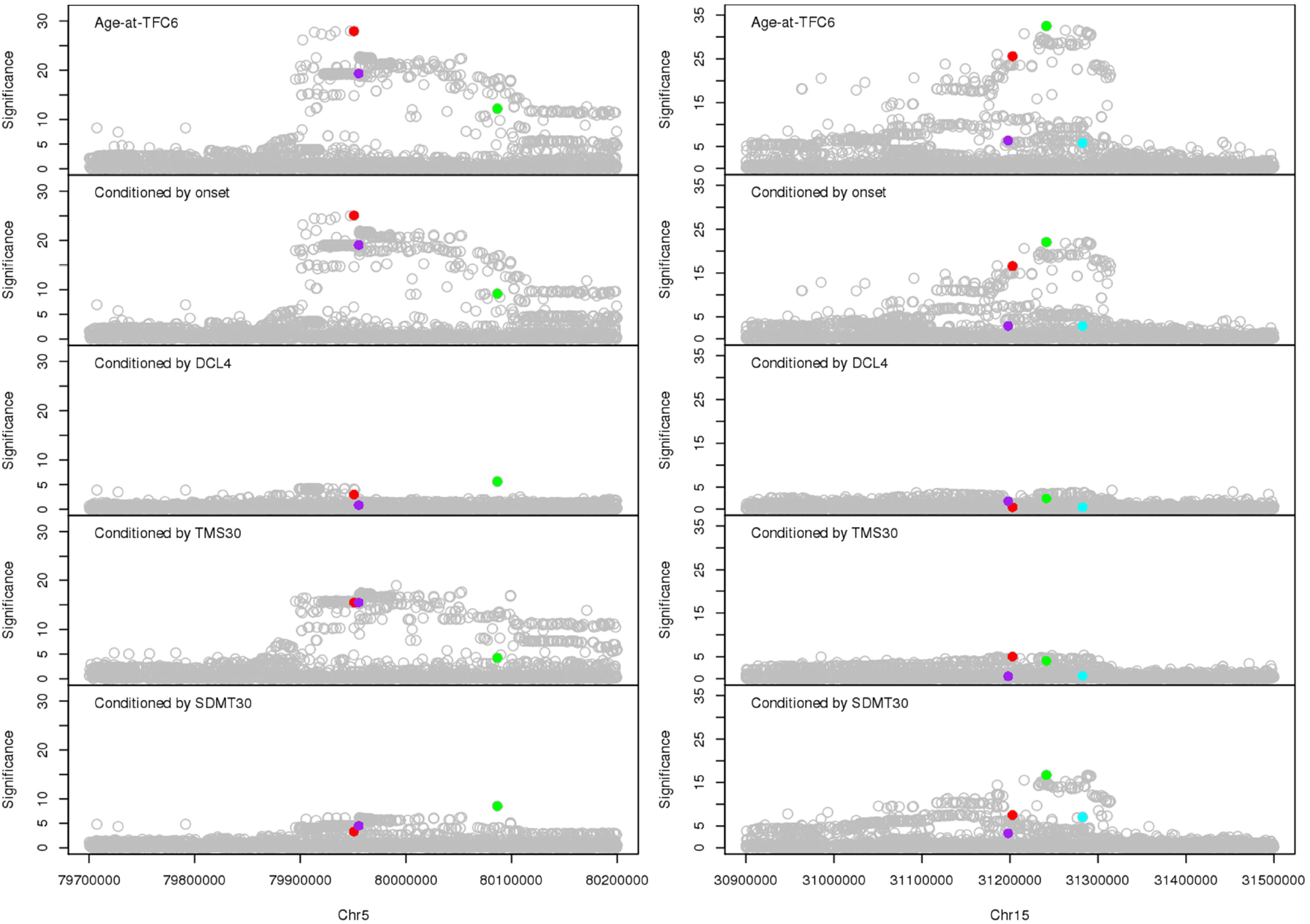
Age-at-TFC6 signal at *MSH3* and *FAN1* conditioned on other phenotypes. Regional plots displaying age-at-TFC6 association significance (-log10(p-value)) at *MSH3* (left, chr 5) and *FAN1* (right, chr 15) loci for the 4,879 participants for whom age-at-onset, age-at-DCL4, age-at-TFC6, age-at-TMS30 and age-at-SDMT30 were all available are shown above equivalent plots where the analysis was conditioned on each of the other age-at-landmark phenotypes. Each circle represents a different SNV with colored symbols indicating tag SNVs for particular modifier effects derived from the analysis of all 6,900 participants with age-at-TFC6 data: 5AM1, 5_79950781_A_G (red), 5AM2, 5_80086504_A_G (green); 5AM3, 5_79955360_G_GT (purple, peak SNV after conditioning on 5AM1 and 5AM2); 15AM1, 15_31202961_G_A (red), 15AM2, 15_31241346_G_A (green); 15AM3, 15_31197995_C_T (purple); 15AM5, 15_31282611_T_C (cyan).

The age-at-TMS30 and age-at-SDMT30 analyses also supported additional complexity of modifier effects at some loci where the top SNVs were not highly correlated across phenotypes. At *MSH3*, the common landmark-hastening 5AM1 modifier effect was captured by highly correlated peak SNVs across all four predicted landmarks (r-square > 0.97) while the rare 5AM2 delaying effect was revealed by the same tag SNV for all phenotypes except age-at-SDMT30, where it did not achieve suggestive significance (Table S1). However, the top common SNVs associated with landmark delay differed for each of the four predicted landmarks, with pairwise r-square values from 0.44 to 0.99, pointing to potential differences in the sources of the modifier effects. Consequently, we first performed association analyses conditioning simultaneously on the 5AM1 and 5AM2 peak SNVs to identify the most significant remaining modification signal for each phenotype. Each landmark yielded a different top SNV tagging a common landmark-delaying effect (Table 2) but only those for age-at-DCL4 and age-at-TFC6 were very highly correlated. We compared separate association analyses across all four predicted landmark phenotypes conditioning each time simultaneously on the 5AM1 and 5AM2 peak SNVs plus one of the top remaining SNVs defined in the previous step. Each of these conditional analyses showed greatly reduced signal for all phenotypes, indicating that the common causative landmark-delaying variants are not independent (Table 2). However, weak to suggestive significant SNVs remained, suggesting the possibility that relative strength of the effect on motor and cognitive circuits differs based on multiple variants in the local haplotype.

**Table 2.**
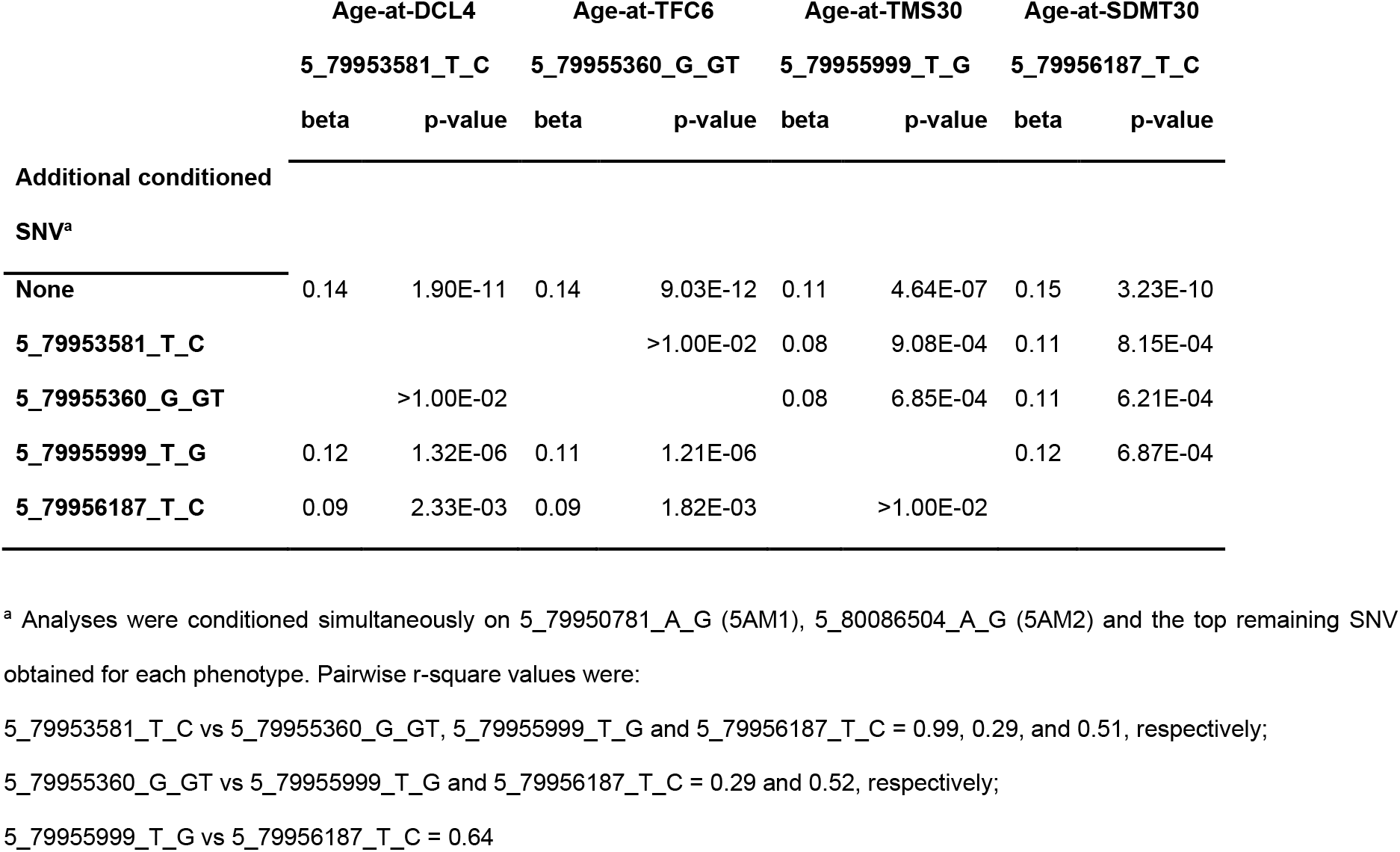
Conditional analysis for common landmark-delaying effects at *MSH3*.

|  | Age-at-DCL4 |  | Age-at-TFC6 |  | Age-at-TMS30 |  | Age-at-SDMT30 |  |
| --- | --- | --- | --- | --- | --- | --- | --- | --- |
|  | 5_79953581_T_C |  | 5_79955360_G_GT |  | 5_79955999_T_G |  | 5_79956187_T_C |  |
|  | beta | p-value | beta | p-value | beta | p-value | beta | p-value |
| **Additional conditioned** |  |  |  |  |  |  |  |  |
| **SNV<sup>a</sup>** |  |  |  |  |  |  |  |  |
| **None** | 0.14 | 1.90E-11 | 0.14 | 9.03E-12 | 0.11 | 4.64E-07 | 0.15 | 3.23E-10 |
| **5_79953581_T_C** |  |  |  | >1.00E-02 | 0.08 | 9.08E-04 | 0.11 | 8.15E-04 |
| **5_79955360_G_GT** |  | >1.00E-02 |  |  | 0.08 | 6.85E-04 | 0.11 | 6.21E-04 |
| **5_79955999_T_G** | 0.12 | 1.32E-06 | 0.11 | 1.21E-06 |  |  | 0.12 | 6.87E-04 |
| **5_79956187_T_C** | 0.09 | 2.33E-03 | 0.09 | 1.82E-03 |  | >1.00E-02 |  |  |

<sup>a</sup> Analyses were conditioned simultaneously on 5\_79950781\_A\_G (5AM1), 5\_80086504\_A\_G (5AM2) and the top remaining SNV obtained for each phenotype. Pairwise r-square values were:
5\_79953581\_T\_C vs 5\_79955360\_G\_GT, 5\_79955999\_T\_G and 5\_79956187\_T\_C = 0.99, 0.29, and 0.51, respectively;
5\_79955360\_G\_GT vs 5\_79955999\_T\_G and 5\_79956187\_T\_C = 0.29 and 0.52, respectively;
5\_79955999\_T\_G vs 5\_79956187\_T\_C = 0.64

Of the DNA maintenance genes other than *MSH3*, age-at-SDMT30 detected only *PMS2* and *FAN1* at genome-wide significance. However, at *PMS2*, age-at-SDMT30 also revealed a second landmark-hastening modifier effect (7AM4), tagged at suggestive significance by a missense SNV (7_6026530_C_T; p.M622I) associated with altered PMS2-MLH1 binding.^39^ Conditioning on either of the *PMS2* landmark-hastening SNVs resulted in increased significance for the other, confirming that they represent different modifier effects (Table S3).

The detection of *MLH1, TCERG1, LIG1* and *RRM2B* at genome-wide significance by age-at-TMS30 but not by age-at-SDMT30 suggests that the respective modifier alleles act preferentially on the motor domain, explaining their capture in the age-at-onset GWAS. However, these findings also suggest that TMS as a measure of motor dysfunction does not provide information identical to the expert rater’s subjective assessment of motor function when determining age-at-onset. In particular, the *CCDC82* locus was robustly detected in continuous analysis by the age-at-TMS30 landmark, yet in the continuous analysis of age-at-onset it achieved only suggestive significance (Table S1).

Notably, based on pure, uninterrupted CAG length, the *HTT* CAA-loss haplotype showed genome-wide significance for both age-at-TMS30 and age-at-SDMT30, consistent with that variation at the site of the CAG repeat influencing both motor and cognitive domains. In contrast, the *HTT* CAACAG-duplication haplotype failed to achieve suggestive significance for either phenotype. The age-at-TMS30 and, especially, the age-at-SDMT30 GWAS also revealed several loci with apparent genome-wide significant association detected only by infrequent alleles (MAF < 1%) (Table S2). However, only one of these alleles achieved even suggestive significance in the age-at-onset, age-at-DCL4 or age-at-TFC6 GWAS suggesting that most or all are likely to result from the coincidence of rare alleles in a small number of phenotypic outliers.

### Gene-wide association and pathway analyses

We also performed gene-wide association analysis across the landmark phenotypes (Table S4). As expected from the single SNV analyses, the genes at several modifier loci, particularly the *MSH3* region, showed increased signal for some or all predicted landmarks compared to age-at-onset. Notably, the age-at-SDMT30 results showed the greatest divergence from age-at-onset in the gene-wide analysis, being far more significant at the *MSH3* locus but much less significant at *MLH1, LIG1, RRM2B*, and *TCERG1*. Pathway analyses based upon these data again implicated mismatch repair, as was the case in our HRC-imputation GWAS (Table S5). Mismatch repair pathways topped the “self-contained” association list for all measures, with significance being greatest for age-at-TFC6. Mismatch repair terms were also most significant (p-value < E-5) in “competitive” analysis of age-at-TFC6 results and nominally significant with the other measures. Also, nucleotide excision repair topped the list in the competitive age-at-SDMT30 analysis, driven largely by *FAN1, LIG1*, and *POLD1*, suggesting that genome-wide significant SNVs may emerge at the latter locus in future expanded GWAS analyses. Interestingly, terms related to regulation autophagy (GO:0016239 “positive regulation of macroautophagy” and GO:0010508 “positive regulation of autophagy”) were also significant in the age-at-TFC6 competitive analysis. Although *HTT* was the most significant individual gene in those gene sets, its exclusion did not dramatically reduce the enrichment p-values (e.g., age-at-TFC6 competitive / self-contained enrichments p-values with *HTT:* GO:0016239 1.40E-07 / 7.64E-07, GO:0010508 4.00E-06 / 3.50E-08; excluding *HTT:* GO:0016239 3.76E-07 / 9.05E-06, GO:0010508 8.44E-06 / 2.8E-07) arguing that the pathway enrichment is due to a number of modestly significant genes rather than a few highly significant genes.

## Discussion

The identification of genetic modifiers (*i.e*., enhancers and suppressors of selected phenotypes) has long been used in model organisms as a route to define the genes and pathways that influence the manifestation of chosen phenotypes, thereby helping to elucidate the molecular mechanisms underlying them. In humans, the discovery of genetic modifiers has the same potential, with the added benefit that for disease phenotypes, this knowledge can potentially target and guide the development of rational therapies. Indeed, the delineation of enhancer and suppressor haplotypes at DNA maintenance genes associated with instability of trinucleotide repeats has pointed to somatic CAG expansion as the rate-determining process for HD motor onset and to the encoded proteins as potential therapeutic targets. Our extension of the GWAS approach to recognized landmarks in the clinical course of HD that incorporate features beyond motor disturbance, combined with our updating to TOPMed genome-wide SNV imputation, have now revealed both commonalities and differences with the previous age-at-onset GWAS findings.

First, the integration of multiple phenotypes collected in standardized tests longitudinally across motor, cognitive, and functional domains captured almost all previously recognized GWAS modifier loci at genome-wide significance, often with greater power than did motor age-at-onset. In many cases, the same top SNV was observed across the different landmarks, but in cases where different top SNVs emerged, they were usually highly correlated and of comparable MAF and location, with some notable exceptions discussed below. Importantly, the increased significance was achieved at a considerably lower sample size, indicating that the landmarks algorithmically predicted from standardized measures generally provide more informative, less subjective measures than the judgment-determined age-at-motor onset assigned by a rater. The reduced sample size compared to previous GWAS resulted from not having the requisite standardized measures on many of the earliest subjects from our age-at-onset GWAS^2^ or sufficient numbers of participants with CAG 51-55 to permit algorithmic prediction in this range. However, between implementation of TOPMed imputation and the algorithmic prediction strategy, we also identified multiple modifier effects at the *PMS1* and *PMS2* loci and potentially a novel onset-delaying locus (7BM1). The latter does not harbor a DNA maintenance gene, suggesting that it might influence the toxicity mechanism precipitated by the somatically expanded CAG repeat. Overall, the success of algorithmic landmark prediction bodes well for significantly expanding the available sample size for HD modifier GWAS by genotyping of all subjects in the natural history studies, regardless of onset status. It also argues for extending the strategy in the future to additional landmarks and phenotypes, both before and after HD clinical diagnosis.

A second notable finding in the age-at-DCL4 and age-at-TFC6 GWAS compared to the motor age-at-onset GWAS is that the level of significance for select modifier loci was increased disproportionally to the rest. This indicates that beyond improved precision of the predicted phenotypes, there is also a difference in the genetic information reflected by the predicted vs. observed landmarks. At *MSH3*, this relative increase was due at least in part to the inclusion of SDMT as a cognitive measure in the cUHDRS, which was used to perform the age-at-DCL4 and age-at-TFC6 landmark predictions. Both analysis of individuals with extreme TMS or SDMT scores and subsequently, of two new predicted landmarks based upon these measures, indicated that the common *MSH3* landmark-hastening and landmark-delaying modifier effects were better captured by SDMT than TMS, while the common onset-delaying *FAN1* effect exhibited the reverse pattern. The differential impact of these two loci on cognitive and motor domains was also evident in association analyses conditioned by phenotype (Figure 5). Thus, although *MSH3* and *FAN1* both influence somatic instability of the *HTT* CAG repeat, their modifier effects differentially impact neuronal circuits critical for motor vs. cognitive dysfunction in HD. Indeed, the common 5AM1 and 15AM2 modifiers are both associated with brain eQTLs (expression quantitative trait loci) for *MSH3* and *FAN1*, respectively, and it is likely that, like risk loci in common disease, these and many of the other HD modifier effects act via regulation of gene expression that can vary across cell types.^3^ The relatively more significant impact on the cognitive domain for the common *MSH3* modifier effects is also consistent with detection of this locus as a modifier of HD progression by TRACK-HD investigators who constructed their GWAS phenotype from the first principal components of longitudinal measures, but specifically excluded TMS.^20^ It has been suggested that the basis for this progression-delaying effect is a particular allele of a functional coding repeat in exon 1 of *MSH3*.^40^ Our data reveal subtle haplotype differences when comparing the common landmark-delaying versions of this locus across phenotypes, potentially pointing to differences in regulation of *MSH3* expression in the context of the same version of the functional repeat in the *MSH3* protein. However, it is not possible to exclude distinct functional variants or an effect on the immediately adjacent/overlapping *DHFR* gene. Of note in our study, among those participants with both predicted-phenotypes, the infrequent landmark-delaying 5AM2 effect showed genome-wide significance for age-at-TMS30 (p-value 1.18E-09) but not even suggestive significance for age-at-SDMT30. This could indicate that, unlike the common landmark-delaying effect, this rarer *MSH3* locus modifier whose mode of action is not yet known acts differentially on cell types/neuronal networks contributing preferentially to the motor phenotype.^41^

At *CCDC82*, which encodes a coiled-coil domain protein of unknown function, the relative increase in signal with age-at-DCL4 and age-at-TFC6 was due at least in part to the role of TMS, not SDMT, in making these landmark predictions. This locus did not achieve genome-wide significance in the continuous age-at-onset GWAS but did in the age-at-TMS30 GWAS (Table S1) and conditional analysis showed that the age-at-TFC6 signal was removed by conditioning on age-at-TMS30, but not on either age-at-onset or age-at-SDMT30 (Figure S5).

At *PMS2*, the increased significance achieved with the age-at-DCL4 and age-at-TFC6 landmarks appears to reflect effects of both TMS and SDMT on the predictions. The onset-delaying 7AM1 effect tagged by SNVs of ∼15% MAF yielded genome-wide significance in the age-at-onset analysis but could not be directly visualized in the age-at-landmark analyses where far more significant p-values were achieved for landmark-delay tagged by SNVs of ∼41% MAF (7AM3). The situation is complicated because analyses conditioning on the respective top SNVs suggest that the haplotype responsible for the 7AM1 effect is a distinct subset of those chromosomes possessing the haplotype of 7AM3-associated SNVs (Table S3). The association signals across landmarks indicate that the 7AM1 subset preferentially influences age-at-onset while the 7AM3 superset (including both 7AM1 and non-7AM1 versions of 7AM3) has a relatively greater influence on the predicted landmarks, suggesting the presence of more than one functional variation on the 7AM1-tagged chromosomes. Conditioning on age-at-onset reduces the age-at-TFC6 signals to a lesser extent than does conditioning on either age-at-TMS30 or age-at-SDMT30 (Figure S6). Despite its clear association with age-at-onset, the 7AM1 effect was not responsible for the peak signal in the age-at-TMS30 analysis, indicating, like the *CCDC82* findings, a difference even within the motor domain between the quality and/or nature of the information captured by TMS versus rater-determined motor onset.

The notion that even phenotypes focused on the same domain can reveal differential detection of modifier loci, indicating that these phenotypes constitute subtly different information, is also reflected in the results for other modifiers that preferentially influence the motor domain. The locations of the DNA mismatch repair gene *MLH1*, the DNA ligase gene *LIG1*, the transcription factor gene *TCERG1* and the ribonucleotide reductase subunit gene *RRM2B* were all captured at genome-wide significance in the age-at-TMS30 GWAS but only *MLH1* and *LIG1* yielded even suggestive significance in the age-at-SDMT30 GWAS (Table S1). All were also captured at genome-wide significance in the age-at-DCL4 and age-at-TFC6 analyses, presumably due mainly to the contribution of TMS to the cUHDRS-based prediction of these landmarks. This supposition is consistent with the preferential reduction in the age-at-TFC6 signal at all four loci when conditioning on age-at-TMS30 compared to age-at-SDMT30 (Figure S7). A difference in the nature of the information provided by TMS and age-at-onset, along with differences in the imputation with TOPMed vs. HRC data, might also be responsible for the detection of the potential new modifier locus on chr 7 (7BM1) only in the age-at-onset GWAS. This locus, which is in the general vicinity of *CHCHD2*, a gene that is associated with Lewy Body disease and was recently suggested from combined gene expression profiling and GWAS as a potential HD onset modifier^42^, remains to be confirmed through an expanded sample size.

Our prior genetic studies have pointed to two potential types of modifiers in HD, those that affect the rate at which somatic expansion of the CAG repeat leads to disease onset and those that influence the toxicity mechanism or its downstream consequences which lead to neuronal dysfunction and death. Modifiers of the first type have already been identified involving DNA maintenance genes implicated by GWAS, but it is not yet certain whether any of the other genome-wide significant GWAS loci fall into the second category. Our pathway analyses and previous cellular and molecular studies suggest that regulation of macroautophagy may play a role,^43; 44^ but confirmed GWAS genome-wide significance would bolster this hypothesis. Taken as a whole, our current results indicate that different HD phenotypes capture the effects of individual genetic modifier haplotypes differentially. By corollary, even modifier alleles that are thought to influence HD via the same molecular process, i.e., somatic CAG expansion, might do so preferentially in different cell types or at different times. This is because if the modifiers effects were on *HTT* CAG length in the same cell types and at the same times, they would be expected to influence the same phenotype(s). The cells in which a modifier effect is felt may depend on such factors as the critical CAG length threshold to trigger toxicity, which may vary by cell type, and also on the mechanism by which the genetic variation acts at the locus. For example, functional variants that act through altering the level of expression of the modifier gene might well do so in a cell type-specific or type-preferential manner based upon differential regulation of expression, whereas variants that alter the protein’s structure have the potential for more global effects. It is important to note, however, that our studies do not argue that the participation of any of the DNA maintenance modifier genes in the process of CAG repeat instability is limited to particular sets of cells. Rather, they indicate that these genes have a differential impact on CAG instability depending on the genetic variations that allow them to be recognized as modifiers. Unravelling the actions of the various modifier effects is likely to require both the consideration of a variety of phenotypes at the human subject level and analysis of functional cell-type differences, including single-cell and molecular analyses. However, empowered by longitudinal natural history studies collecting standardized clinical measures, such expanded genetic analyses offer great promise for dissecting the complexity of the HD disease process and developing the most effective treatments.

## Data Availability

Original data will be made available post-publication on request. Data involving human subjects will be shared with qualified investigators given their institutional assurance of subject confidentiality and compliance with GDPR requirements with respect to personal data. Requests for further information or for other resources and reagents should be directed to James F. Gusella, Ph.D.

## Supplemental data

Supplemental data include seven figures and five tables (in 1 excel file).

## Acknowledgments

These HD studies would not be possible without the vital contribution of the research participants and their families. Individuals who contributed to the collection subject data can be found at https://www.enroll-hd.org/enrollhd_documents/2018-10-R1/Enroll-HD-Acknowledgement-List-Public-2018-10-R1.pdf, in the Supplementary Material of Lee et al. (2017), and in the Supplemental Information of Genetic Modifiers of Huntington’s Disease (GeM-HD) Consortium (2015). Supported by the CHDI Foundation (U.S.), the U.S. National Institutes of Health (NS082079, NS091161, NS016367, NS049206, NS105709, NS114065, NS119471, NS040068), the Medical Research Council (UK; MR/L010305/1 and fellowship MR/P001629/1), and a Cardiff University School of Medicine studentship.

## Web resources

ENROLL-HD platform, https://www.enroll-hd.org/

European Huntington’s Disease Network, http://www.ehdn.org/

GEMMA program, https://github.com/genetics-statistics/GEMMA

GenABEL program, https://cran.r-project.org/src/contrib/Archive/GenABEL/

Gene Ontology website, http://geneontology.org/docs/download-ontology/

Huntington Study Group, https://huntingtonstudygroup.org

HRC or 1000G Imputation preparation and checking program, https://www.well.ox.ac.uk/∼wrayner/tools/

MAGMA v1.08, https://ctg.cncr.nl/software/magma

Molecular Signatures Database, https://www.gsea-msigdb.org/gsea/msigdb/index.jsp

NCBI, https://ftp.ncbi.nlm.nih.gov/gene/DATA/

OMIM, http://www.omim.org/

R package flexsurv, https://cran.r-project.org/web/packages/flexsurv/index.html

R package lme4, https://cran.r-project.org/web/packages/lme4/index.html

R package lqmm, https://cran.r-project.org/web/packages/lqmm/index.html

Reactome, https://reactome.org/download-data

TOPMed Imputation Server, https://topmedimpute.readthedocs.io/en/latest/

