## Supplemental Data for "Genetic modifiers of Huntington’s disease differentially influence motor and cognitive domains"

#### Figure S1 – Algorithmic prediction of age-at-landmark HD phenotypes

Phenotype algorithm for estimated age-at-DCL4 (and TFC6). (1) Parametric survival curve (Weibull regression) with left-truncation and right-censoring is estimated for each CAG/sex/education (low= high school or less, high= more than high school) combination and the median age (A) at survival is computed (survival from conversion to DCL4). (2) Linear mixed model (LMM) with cubic splines is used to estimate the mean cUHDRS trajectory and the value at which the mean trajectory intersects with A is determined, which is the cUHDRS threshold (cUHDRS = 15 in this example). (3) Individual cUHDRS trajectories are estimated based on the LMM for observed age (solid line) and extrapolated over the entire age range (dotted line). The age at which an individual's predicted curve intersects with the cUHDRS threshold is the estimated age at the landmark (ai). (4) Each estimated age (ai) is transformed to a Z-score by correcting for the median age (A) and scaling by the standard deviation (SD) of all the ages for the individuals in the CAG/sex/education cohort (which adjusts for these variables). A similar method is used for estimating age-at-TFC6.

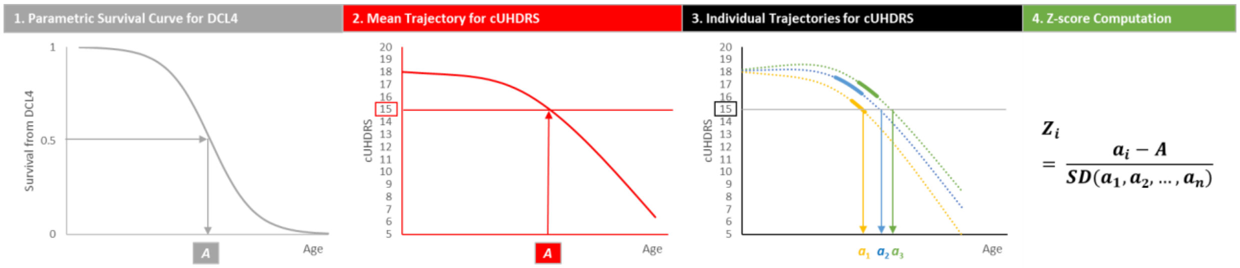

**Figure S2 – Comparison of continuous age-at-onset GWAS with genotypes imputed with TOPMed or Haplotype Reference Consortium data**

Stacked plots comparing continuous age-at-onset GWAS using genotypes imputed using TOPMed data (top) or Haplotype Reference Consortium data (bottom) for 9,009 HD individuals (the subset of the 9,058 individuals from our published GWAS3 who remained after quality control of the TOPMed imputation data). Arrows indicate loci that yield genome-wide significant signal in the TOPMed analysis (excluding signals from single rare variants (< 1% MAF) except at *HTT*). Implicated modifier genes are labeled in the top panel. Red labels indicate DNA maintenance genes. One novel locus, described in the text, is labeled by its HD modifier effect designation (7BM1) based upon our standard nomenclature and a dashed arrow.

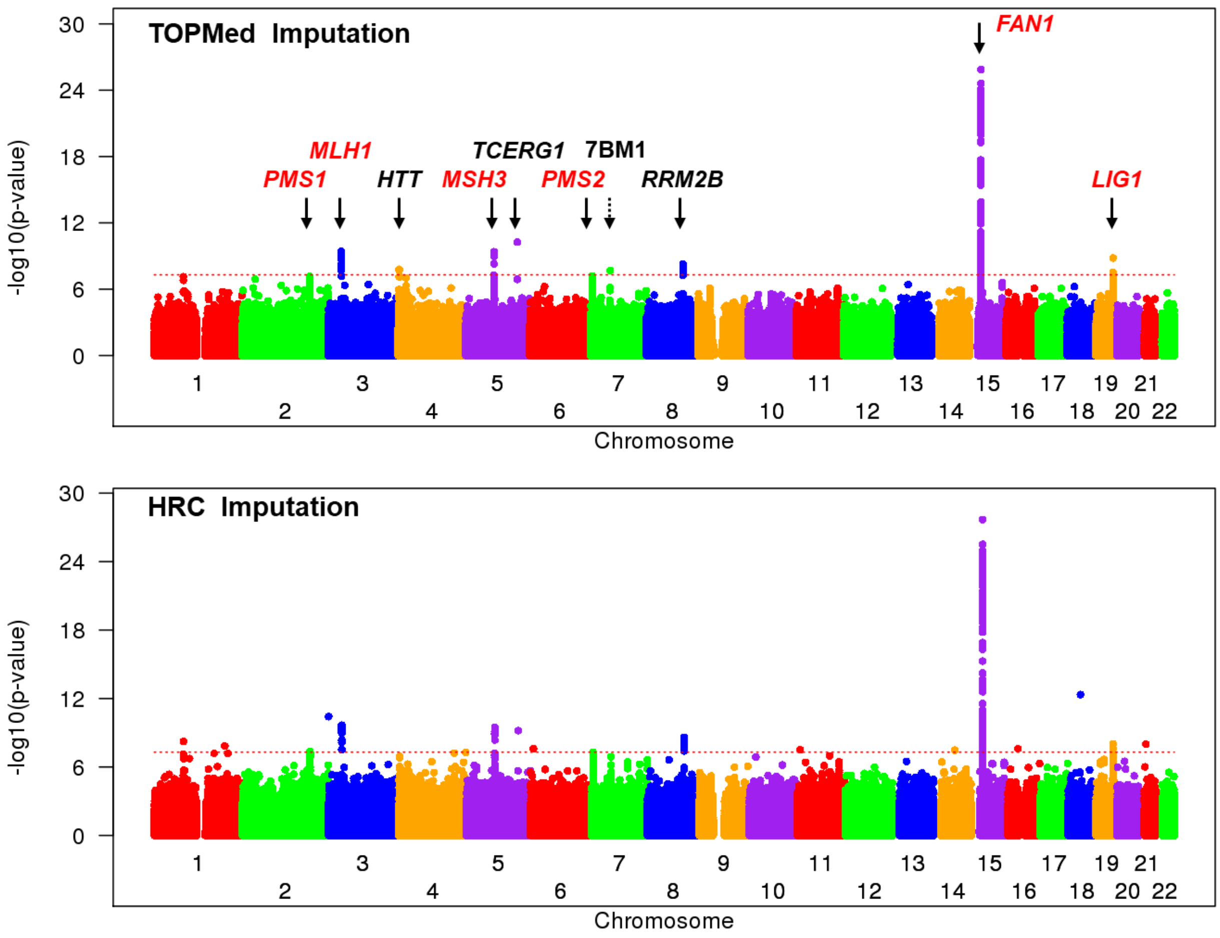

**Figure S3 – Distribution of inherited CAG repeat lengths and ages-at-onset in the study sample**

A histogram of the number of HD subjects in our study sample with inherited CAG repeat lengths from 40 to 55 is shown at the left.  At the right is a standard box plot showing the median and range of ages at onset by CAG repeat length, indicating a wider variation at lower inherited CAG repeat lengths. For each CAG repeat, the top, middle, and bottom of the box represent 75th percentile (upper quartile), median, and 25th percentile (lower quartile) data points, respectively. The higher and lower whiskers represent the upper quartile +1.5 X interquartile range (IQR) and the lower quartile –1.5 X IQR, respectively. Circles (i.e., outliers) represent data points outside of the whiskers for a given CAG repeat.

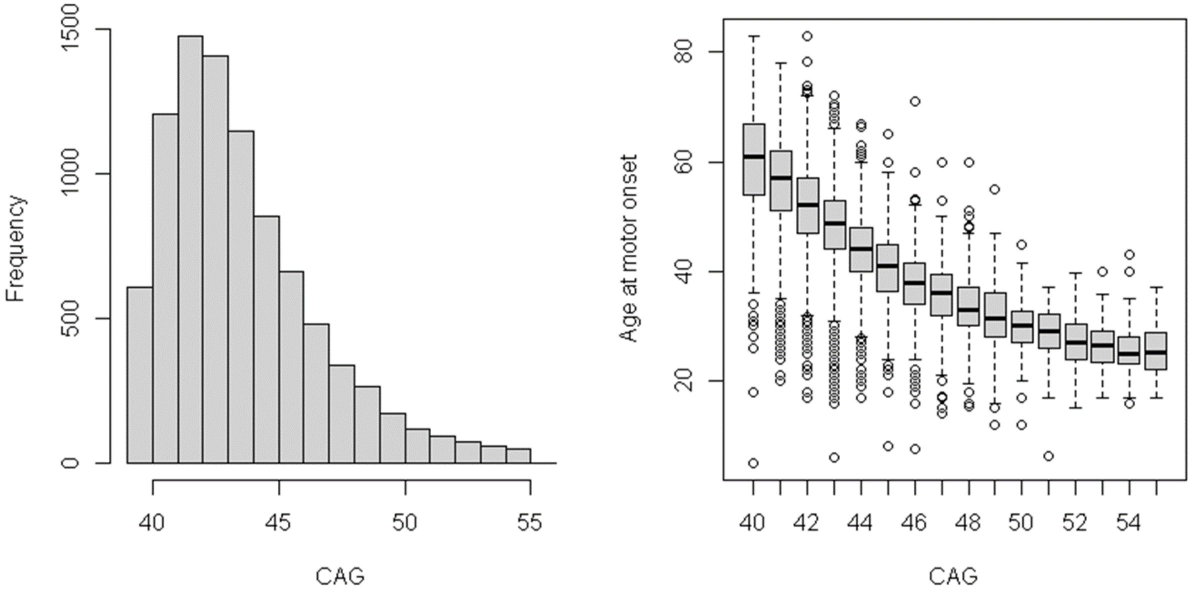

**Figure S4 – Regional association plot indicating tag SNVs for haplotypes bearing CAA-loss and CAACAG-duplication variants in *HTT***

GWAS signals in the *HTT* region are shown after correction of CAA-loss and CAACAG-duplication SNV haplotype individuals for pure CAG length. Variants are designated by triangles whose size reflects the frequency of the minor allele and orientation indicates the direction of effect for that minor allele (upward-pointing, onset-delaying; downward-pointing, onset-hastening). Subjects bearing CAA-loss and CAACAG-duplication variants were implicated by the tag SNVs indicated by orange (CAA-loss) and green (CAACAG-duplication) arrows, respectively, and the presence of the variant and uninterrupted CAG length was confirmed by DNA sequencing. Location of the *HTT* CAG repeat is denoted by the dashed blue line.

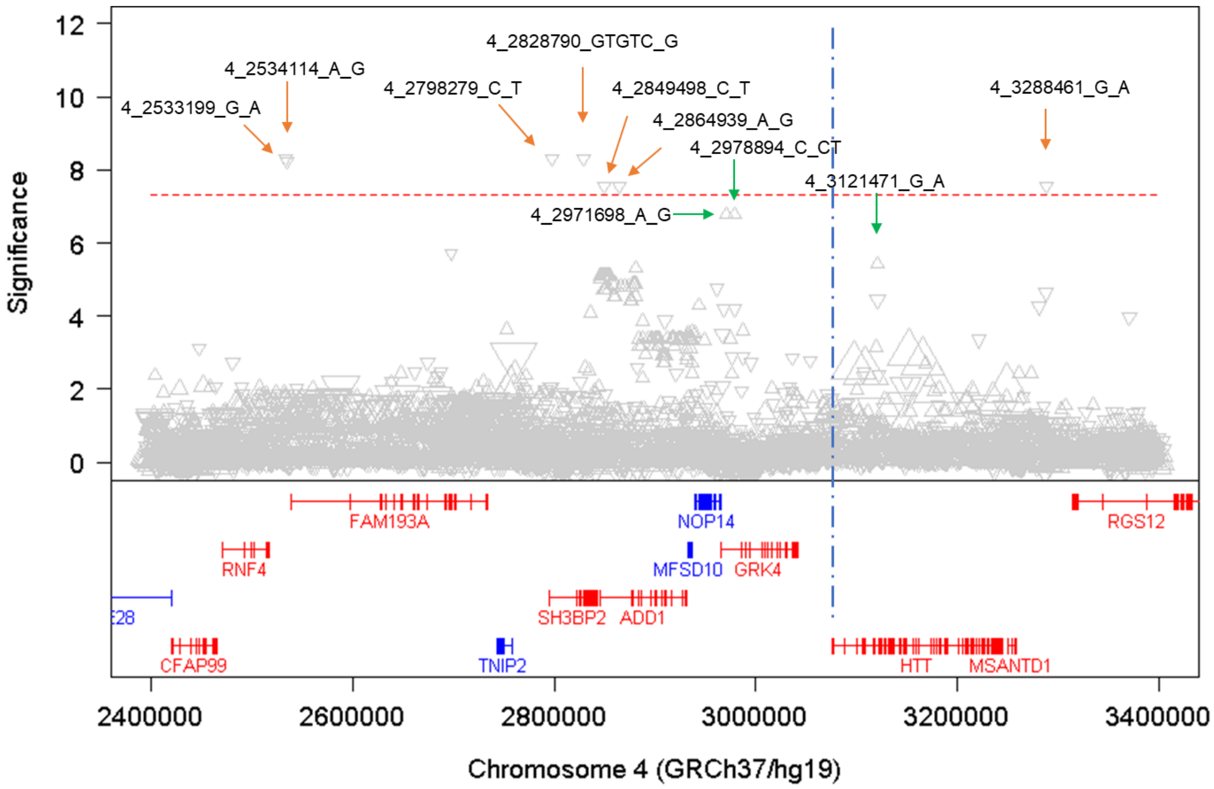

**Figure S5 – Age-at-TFC6 signal at *CCDC82* conditioned on other phenotypes**

A regional plot displaying age-at-TFC6 association results at *CCDC82* (chr 11) for the 4,879 participants for whom age-at-onset, age-at-DCL4, age-at-TFC6, age-at-TMS30 and age-at-SDMT30 were all available is shown above equivalent plots where the analysis was conditioned on each of the other phenotypes. Each circle represents a different SNV with the red symbol indicating the 11AM1 tag SNV (11_96084542_A_C) from analysis of all 6,900 participants with age-at-TFC6 data.

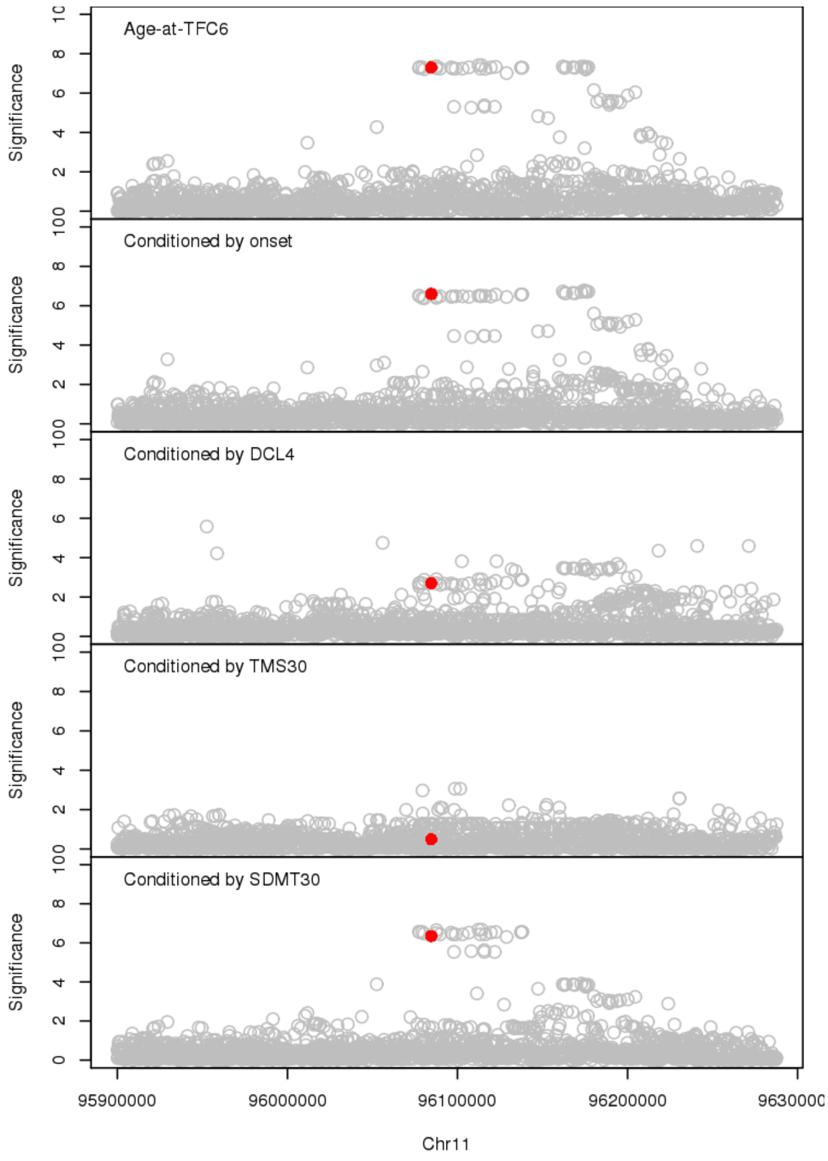

**Figure S6 – Age-at-TFC6 signal at *PMS2* conditioned on other phenotypes**

A regional plot displaying age-at-TFC6 association results at *PMS2* (chr 7) for the 4,879 participants for whom age-at-onset, age-at-DCL4, age-at-TFC6, age-at-TMS30 and age-at-SDMT30 were all available is shown above equivalent plots where the analysis was conditioned on each of the other phenotypes. Each circle represents a different SNV with colored symbols indicating peak SNVs for particular modifier effects defined the full set of participants with age-at-onset, 7AM1, 7_6022626_C_T (red), age-at-TFC6, 7AM2, 7_6056484_G_C (green) and 7AM3, 7_6041836_T_A (purple) and age-at-SDMT30, 7AM4, 7_6026530_C_T (gold).

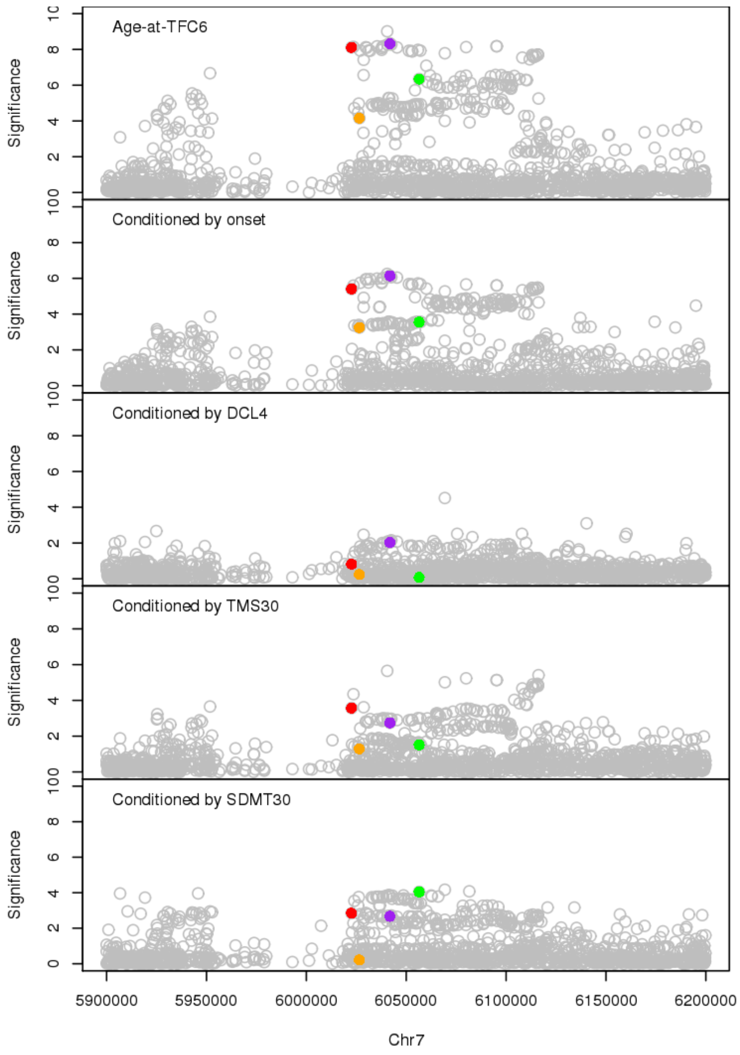

**Figure S7 – Age-at-TFC6 signal at *MLH1*, *TCERG1*, *RRM2B* and *LIG1*, conditioned on other phenotypes**

Regional plots displaying age-at-TFC6 association results at *MLH1* (chr 3), *TCERG1* (chr 5), *RRM2B* (chr 8) and *LIG1* (chr 19) loci for the 4,879 participants for whom age-at-onset, age-at-DCL4, age-at-TFC6, age-at-TMS30 and age-at-SDMT30 were all available are shown above equivalent plots where the analysis was conditioned on each of the other phenotypes. Each circle represents a different SNV with the colored symbols indicating those frequent SNVs (> 1% MAF) that displayed peak signal in the analysis of all 6,900 participants with age-at-TFC6 data: 3AM1, 3_37121844_G_A (red, first panel); 5BM1, 5_145886836_G_A (red second panel); 8AM1, 8_103213640_G_T (red third panel); 19AM1, 19_48683973_C_T (red, fourth panel); 19AM2, 19_48639235_G_A (green, fourth panel).

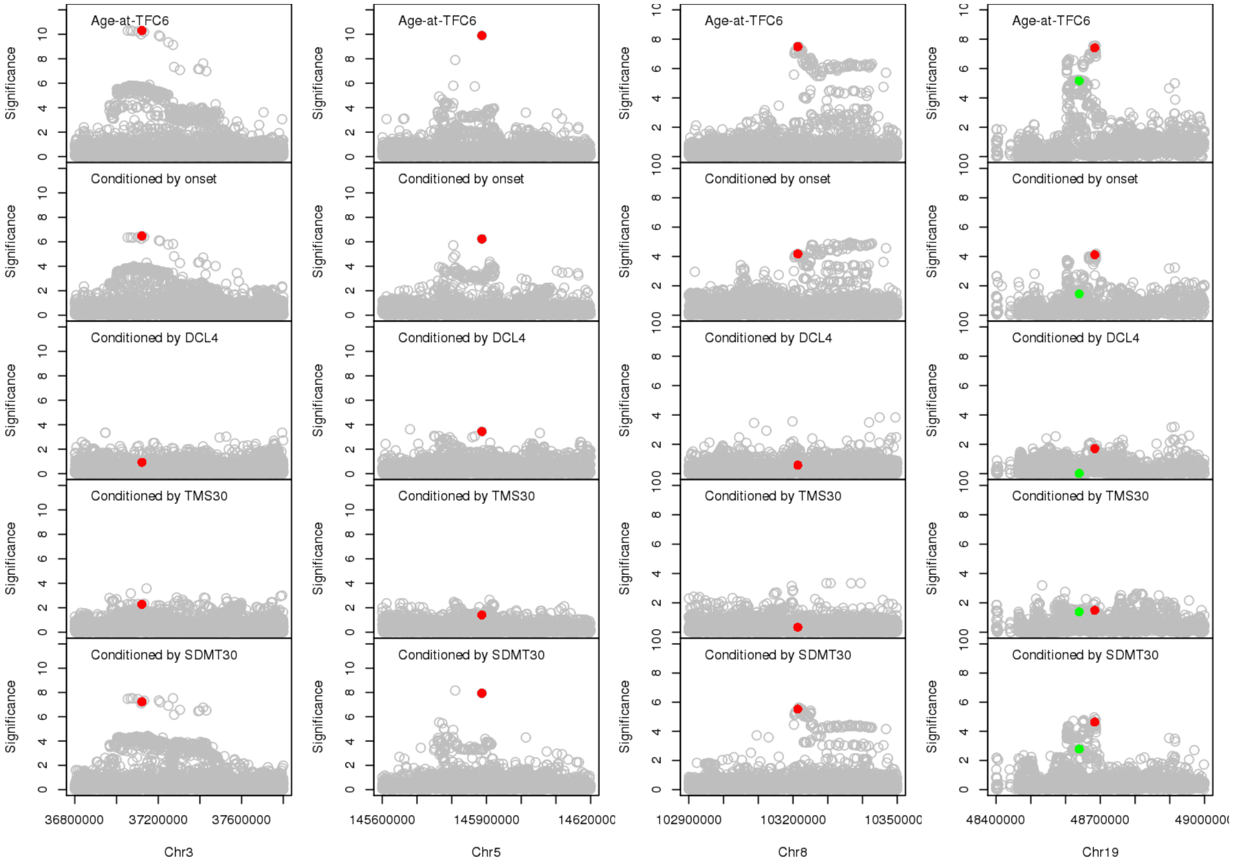

### Supplemental Tables

**Table S1 - Peak P-values for variations tagging implicated HD modifier effects**

**Table S2 - Genome-wide significant signals from SNVs with minor allele frequency < 1%**

**Table S3 – Conditional analysis for *PMS2* region haplotype tag variants**

**Table S4 – Gene-wide association analyses**

**Table S5 – Pathway analyses**

See excel file Supplemental_Tables.xls
